# Dominant role of abdominal adiposity in circulating lipoprotein, lipid, and metabolite levels in UK Biobank: Mendelian randomization study

**DOI:** 10.1101/2021.05.29.21258044

**Authors:** Joshua A. Bell, Tom G. Richardson, Qin Wang, Eleanor Sanderson, Tom Palmer, Venexia Walker, Linda M. O’Keeffe, Nicholas J. Timpson, Anna Cichonska, Heli Julkunen, Peter Würtz, Michael V. Holmes, George Davey Smith

**Author notes:** **Corresponding author:** Joshua A. Bell, Oakfield House, University of Bristol, UK, BS8 2BN, Twitter: @joshuaa_bell. Joint senior authors (Michael V. Holmes, George Davey Smith).

## Abstract

**Background:** The causal impact of excess adiposity on systemic metabolism is unclear. We used multivariable Mendelian randomization to compare the direct effects of total adiposity (using body mass index (BMI)) and abdominal adiposity (using waist-to-hip-ratio (WHR)) on circulating lipoproteins, lipids, and metabolites with a five-fold increase in sample size over previous studies.

**Methods:** We used new metabolic data on 109,532 UK Biobank participants. BMI and WHR were measured in 2006-2010, during which EDTA plasma was collected. Plasma samples were used in 2019-2020 to quantify 249 metabolic traits with high-throughput nuclear magnetic resonance spectroscopy including subclass-specific lipoprotein concentrations, apolipoprotein B, cholesterol and triglycerides, plus pre-glycemic and inflammatory metabolites. We used two-stage least squares regression models with genetic risk scores for BMI and WHR as instruments to estimate the total (unadjusted) and direct (mutually adjusted) effects of BMI and WHR on metabolic traits. We also estimated the effects of BMI and WHR on statin use, and examined interaction of main effects by sex, statin use, and age as a proxy for medication use.

**Results:** Higher BMI (per standard deviation (SD) or 4.8 kg/m^2^) was estimated to moderately decrease apolipoprotein B and low-density lipoprotein (LDL) cholesterol before and after adjustment for WHR, whilst higher BMI increased triglycerides before but not after WHR adjustment. Estimated effects of higher WHR (per SD, or 0.090 ratio-unit) on lipoproteins, lipids, and metabolites were often larger than those of BMI, but null for LDL cholesterol, and attenuations were minimal upon adjustment for BMI. Patterns of effect estimates differed by sex, e.g., only BMI independently increased triglycerides among men, whereas only WHR independently increased triglycerides among women. Higher BMI and WHR (per SD) were each estimated to directly increase the relative odds of using statins (by 3.49 (95% CI = 3.42, 3.57) times higher for WHR). These patterns were most pronounced among women, and there was strong evidence that the effects of BMI and WHR on metabolic traits differed by statin use and age. Among the youngest adults (38-53 years, statin use 5%), higher BMI and WHR (per SD) each modestly increased LDL cholesterol (0.04 SD, 95% CI = -0.01, 0.08 for total effect of BMI and 0.10 SD, 95% CI = 0.02, 0.17 for total effect of WHR). This estimate for BMI fully attenuated, and the estimate for WHR remained unchanged, upon mutual adjustment. These direct effects on LDL cholesterol were more inverse for BMI and less positive for WHR at intermediate ages (54-62 years, statins 17%) and older ages (63-73 years, statins 29%) where the mutually adjusted effects of BMI and WHR on LDL cholesterol had reversed to -0.19 SD (95% CI = -0.27, -0.11) and -0.05 SD (95% CI = -0.16, 0.06), respectively.

**Conclusions:** Our results suggest that abdominal adiposity has a dominant role in driving the metabolic harms of excess adiposity, particularly among women. Our findings also suggest that apparent effects of adiposity on lowering LDL cholesterol are explained by an effect of adiposity on statin use.

## Introduction

Obesity rates have tripled since the 1970s in many high-income countries (1, 2). This is a major public health concern because obesity likely causes the onset of numerous diseases including coronary heart disease (CHD), based on evidence triangulated from human biology plus conventional observational and Mendelian randomization (MR) studies (3-8). Once established, obesity is difficult to reverse via lifestyle (9, 10). It is therefore likely that the direct pharmacological targeting of causal factors that potentially lay between obesity and disease, such as low-density lipoprotein (LDL) cholesterol which raises CHD risk (11-15), will become an increasingly dominant strategy for disease prevention in the coming decades (16-18).

The causal impact of excess adiposity on systemic metabolism – a gateway to numerous diseases including CHD – is unclear. Conventional observational and early MR evidence supports an effect of total adiposity, measured using body mass index (BMI), on raising levels of fasting insulin and glucose (6, 19, 20), plus numerous supporting traits including amino acids, fatty acids, and inflammatory glycoprotein acetyls (GlycA) measured using high-throughput metabolic profiling (19, 21, 22). Total adiposity also likely raises triglyceride levels, but only adipose tissue that is stored abdominally, measured using waist-to-hip ratio (WHR), appears to additionally raise LDL cholesterol (the target trait of statins (13)) (6, 19, 21, 23, 24). Population-based sample sizes for comprehensive metabolic profiling have so far been modest (N < 25,000). Whether abdominal adiposity has a dominant role in raising LDL cholesterol and related traits independent of total adiposity, or whether the total volume of adiposity is what predominantly drives these effects, is an open question.

Conventional observational studies suggest that BMI, WHR, waist circumference (WC), and trunk fat mass index from dual-energy x-ray absorptiometry (DXA) scans all generate comparable estimates of effect on lipid, glycemic, and inflammatory trait levels (6, 21, 25). Mutual adjustment of adiposity measures to estimate the direct/isolated effect of each, i.e., the effect which does not operate via the other feature of adiposity being considered, is problematic, however, because of their co-dependence. Such adjustments conventionally, e.g., when statistically adjusting BMI for WHR, are akin to conditioning on a likely mediator and could induce so-called collider bias via confounders of the mediator-outcome effect, yielding unreliable results (26-28). This is in addition to potential bias from residual confounding in observational studies by confounding factors which are unmeasured or poorly measured. Multivariable MR, an instrumental variable (IV) approach, should be less prone to confounding and collider bias because of its use of randomly allocated instruments and predicted rather than measured exposure values (29). Multivariable MR has not yet been applied to BMI and WHR to estimate the direct/isolated effect of each on an extensive metabolic trait profile, but this could help reveal the dominant causal driver of metabolic disease susceptibility.

In this study, we aimed to compare the direct effects of total and abdominal adiposity on markers of systemic metabolism, including non-HDL lipids and a set of supporting traits which are indicative of glycemic and inflammatory activity. We used new high-throughput metabolic data on ∼110,000 adults from the UK Biobank study to compare MR estimates of the direct effects of BMI and WHR, in addition to conventional observational estimates of effect, together enabling more robust causal inference. We examined whether effects differ importantly by sex, given conventional observational evidence of a more adverse metabolic profile of total and abdominal adiposity among women in middle age (30). We also examined whether adiposity influences the use of statins, given that statins are commonly prescribed in middle age and they alter non-HDL lipids and metabolites (13, 31). Because this could distort the effects of adiposity on metabolic traits, we examined whether effects differ importantly by statin use and by age as a proxy for mediation use.

## Methods

### Study population

UK Biobank is a prospective cohort study in which 502,549 adults aged 40-69 years were recruited between 2006-2010 via 22 assessment centres across England, Wales, and Scotland (32). This sample represents ∼5% of the 9.2 million adults registered with the UK National Health Service who were invited to participate; potential biases from low representativeness are discussed elsewhere (33, 34) and in current limitations. Full descriptions of the study design, participants, and quality control (QC) are published (35). Participants provided written informed consent. Ethical approval was obtained from the North West Multi-centre Research Ethics Committee (11/NW/0382). Data were accessed via application numbers 30418 and 15825.

Nearly all participants provided blood samples at the 2006-2010 assessment centre for genotyping and biochemistry analyses. Genotype was measured from serum samples using a genome-wide array (UK Biobank Axiom Array) with imputation to the Haplotype Reference Consortium panel. Pre-imputation QC, phasing, and imputation are described elsewhere (36). Our analyses were restricted to autosomal variants using graded filtering with varying imputation quality for different allele frequency ranges (37). 814 individuals with a mismatch between genetic and reported sex, and with sex-chromosome aneuploidy, were excluded. We further restricted to individuals of ‘European’ ancestry as defined by k-means clustering using the first 4 principal components (PCs) provided by UK Biobank (37). We included the largest cluster from this analysis (n=464,708). We further restricted our sample to those self-reporting as white British (3,130 excluded) to further reduce potential for confounding by population structure, leaving 461,578 genotyped participants for consideration.

### Exposures and instruments: BMI and WHR

At the 2006-2010 assessment centre, standing height was measured without shoes using a Seca 202 device and body weight was measured using the Tanita BC-418 MA body composition analyzer with heavy outer clothing additionally removed. A non-stretchable Wessex tape measure was used to record waist circumference at the umbilicus and the circumference of the hip, both in cm. BMI was derived as weight in kg divided by height in squared meters. WHR was derived as waist circumference divided by hip circumference.

Genetic risk scores (GRSs) were constructed for BMI and WHR (unadjusted for BMI) using single nucleotide polymorphisms (SNPs) that were independently associated with each exposure (at R^2^<0.001 and P<5×10^−8^) in a genome-wide association study (GWAS) meta-analysis of between 221,863 and 806,810 adults of European ancestry from the Genetic Investigation of ANthropometric Traits (GIANT) consortium and UK Biobank (38). Separate sets of SNPs were used based on sex-combined and sex-specific GWAS (outlined in **Table 1**). GRSs were constructed using PLINK 2.0, with GWAS effect alleles and beta coefficients as weightings. Standard scoring was applied by multiplying the effect allele count (or probabilities if imputed) at each SNP (values 0, 1, or 2) by its weighting, summing these, and dividing by the total number of SNPs used. The score therefore reflects the average per-SNP effect on the exposure. Estimates from separate two-stage least squares models with total cholesterol as an example metabolite outcome among participants eligible for ≥ 1 analysis indicated that each GRS was strongly associated with its respective exposure measured in UK Biobank, particularly among women, with F-statistics from unadjusted and adjusted models far exceeding recommended minimum levels of 10 (39) (**Table 1**). These high F-statistics indicate low potential for ‘winner’s curse’ and weak instrument bias from UK Biobank having been included in both the GWAS and applied MR samples (i.e., sample overlap). There was a modest positive correlation between the BMI GRS and WHR GRS among sexes combined (Pearson r = 0.25). This correlation was larger among men (r = 0.35) than women (r = 0.15) based on sex specific GRSs.

**Table 1.**
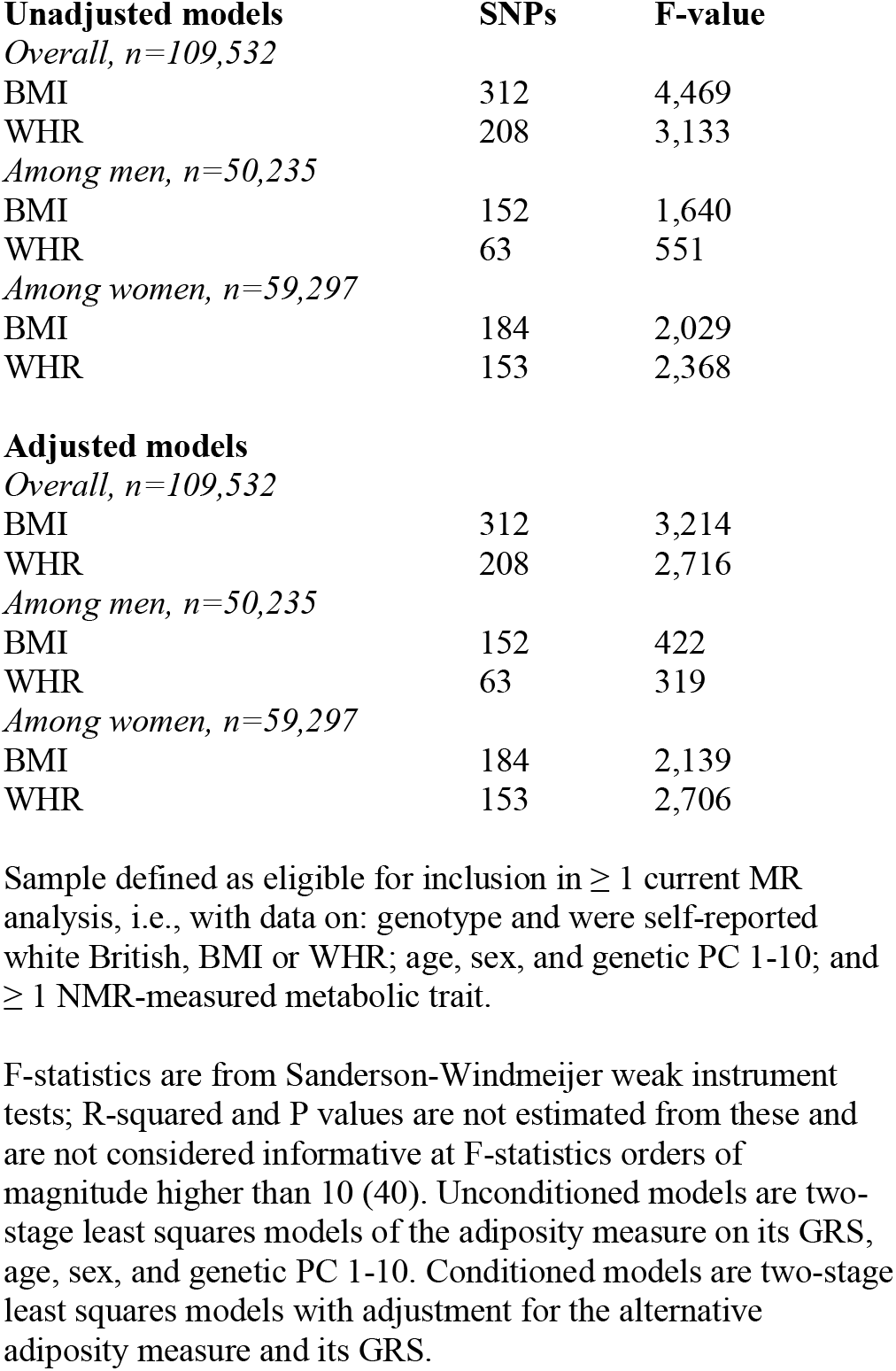
Estimated strength of genetic instruments among participants eligible for ≥ 1 current MR analysis in UK Biobank

### Outcomes: lipoproteins, lipids, and metabolites

EDTA plasma samples from 117,121 participants, a random subset of the original ∼500,000 who provided samples at the 2006-2010 assessment centre (N=113,474) or at a subsequent assessment centre in 2012-2013 (N=3,647), were additionally analysed in 2019-2020 for levels of 249 metabolic traits (168 concentrations plus 81 ratios) using high-throughput proton nuclear magnetic resonance (^1^H-NMR) spectroscopy (data pre-processing and QC steps are described previously (22, 41). These included the cholesterol and triglyceride content of various subclasses of high-density lipoprotein (HDL), LDL (size-specific direct measure and Friedewald-equivalent/clinical direct measure), intermediate density lipoprotein (IDL), and very-low density lipoprotein (VLDL) particles, plus the concentrations and diameter/size of these particles; apolipoprotein B (apoB) and apolipoprotein A-1 (apoA-1) concentrations; in addition to various classes of fatty acids and their ratios to total fatty acid concentration, branched chain and aromatic amino acids, glucose and pre-glycemic factors including lactate and citrate, fluid balance factors including albumin and creatinine, and inflammatory GlycA.

We additionally examined several metabolic traits that were measured using conventional biochemistry (non-NMR) on serum samples from the 2006-2010 assessment centre from the wider UK Biobank participant group for the purpose of comparing effect estimates for those same traits when derived using the NMR platform. These included total cholesterol, LDL cholesterol, HDL cholesterol, total triglycerides, apoB, apoA-1, glucose, albumin, and creatinine. If values from the first assessment centre were missing but values from the subsequent 2012-2013 assessment centre were not missing, then values from this second occasion were used (between 1,206 and 2,199 missing values replaced across traits).

109,532 participants were considered eligible for inclusion in ≥ 1 current MR analysis because they had data on: genotype and were self-reported white British; BMI or WHR; age, sex, and genetic PC 1-10; and ≥ 1 NMR-measured metabolic trait. Analyses were conducted on non-complete-case samples (N varying between models and across traits) to make full use of measured data.

### Main analyses: MR estimates of effect

Exposures and outcomes were internally standardised using z-scores (by subtracting the mean and dividing by the standard deviation, among sexes combined) to enable the comparison of effect sizes given different measurement units. To estimate the total effects of BMI and WHR on each metabolic outcome, separate two-stage least squares (2SLS) regression models were used, with robust standard errors (to accommodate potentially skewed outcome distributions), adjusting for sex, age, and genetic PCs 1-10. In the first stage of these models, the exposure is regressed on its GRS and covariates, and in the second stage, the outcome trait is regressed on the predicted values of exposure and the actual values of covariates from this first stage regression. To estimate the direct effects of BMI and WHR on each metabolic outcome, these 2SLS models were additionally adjusted for the other adiposity measure and its GRS instrument, such that, e.g., BMI is instrumented by the BMI GRS as well as the WHR GRS (plus covariates), and those predicted values are then used in second stage models in relation to outcomes; and likewise, for WHR. Initial evidence for interaction of BMI and WHR with sex in relation to outcomes was tested via P-values for product terms included as covariates in 2SLS models. Main models were then repeated among men and women separately, using sex-specific GRSs for BMI and WHR.

We estimated the prevalence of statin use based on medication codes for self-reported use of any of 13 drugs (atorvastatin, crestor, eptastatin, fluvastatin, lescol, lipitor, lipostat, pravastatin, rosuvastatin, simvador, simvastatin, zocor, zocor heart-pro) as defined in previous genetic analyses of UK Biobank (42). We used this composite statin variable (yes/no) to estimate the effects of BMI and WHR on statin use using 2SLS models with logistic regression as the second stage, and evidence for interaction of BMI and WHR with statin use via product terms of genetically predicted BMI or WHR with observed statin use in relation to outcome traits, expecting interactions to be most evident for LDL cholesterol. We planned sensitivity analyses whereby 2SLS models were repeated with statin users excluded to re-estimate the total and direct effects of BMI and WHR on metabolic outcomes. This was considered preferable to applying an adjustment factor to LDL cholesterol concentration because numerous LDL-related metabolites are examined (not one summary measure), and this enables a uniform approach across lipid subclasses and comparisons with HDL lipids which should be less altered by stratification. To further examine the potential for statins to distort estimated effects of BMI and WHR on metabolic traits, we tested interaction of BMI and WHR with age in relation to outcomes and repeated analyses with stratification by age tertile (instead of statin use). Use of statins and other medications increases markedly with age and estimates at younger ages are expected to be less distorted by medication use. Age is also not influenced by adiposity and so stratifying on age should be less prone to collider bias than stratifying on statin use itself.

### Additional analyses: conventional observational estimates of effect

For comparison with MR results, we conducted conventional observational analyses using multivariable linear regression models with robust standard errors to examine associations of BMI and WHR with differences in the same 249 metabolic traits (plus non-NMR measures for comparison). These models initially adjusted for sex, age at assessment, highest educational qualification, smoking status (‘never’, ‘former’, or ‘current’), and alcohol frequency (‘never’, ‘former’, ‘special occasions only’, ‘once/month to twice/week’, or ‘thrice/week to daily’). Models were then additionally adjusted for the alternative adiposity measure (WHR or BMI) to examine attenuation of BMI and WHR upon mutual adjustment. As done for MR, initial observational evidence for interaction of BMI and WHR with sex in relation to outcome traits was tested via P-values for product terms in linear regression models; models were then repeated among men and women separately. The same interaction tests of BMI and WHR with statin use and age, and the same model stratifications, were conducted as prior.

Because MR models estimate the effects of exposures on outcomes using genetically predicted exposure values, under standard IV assumptions (29, 43), we describe MR estimates in results text using causal language (‘association’ is not considered an appropriate term because models are based on predicted effects, not directly associated variable values). Conventional (multivariable regression) models are based on direct tests of association between variable values, under standard regression assumptions, and we use causal language to describe these estimates in results text for consistency with MR analyses. Our aims involve effect estimation in each instance, and so we focus our interpretations of results on the direction, magnitude, and precision of estimates and provide exact P-values, as recommended (44, 45). Analyses were conducted using Stata 16 (StataCorp, College Station, Texas, USA).

## Results

### Sample characteristics

Eligible participants had a mean (SD) age of 56.7 (8.0) years, which was similar between sexes (**Table 2**). A college or university education was common, at 39.1%, and slightly more so among men. 53.9% reported never having smoked, and this was more common among women than men (58.5% vs 48.5%, respectively). Current smoking was more common among men (12.2% vs 9.0%, respectively). Nearly half reported drinking alcohol at least thrice/week, this again was more common among men. Overall, mean (SD) BMI was 27.4 kg/m^2^ (4.8) which was similar between the sexes, and mean (SD) WHR was 0.87 (0.090) which was higher among men than women (at 0.94 (0.065) and 0.82 (0.069), respectively). The overall prevalence of stain use was 16.4%; this was twice as common among men than women (21.8% vs 11.9%, respectively).

**Table 2.** Characteristics of participants eligible for ≥ 1 current MR analysis in UK Biobank

|  | **Overall**<br>N=109,532 | **Men**<br>N=50,235 | **Women**<br>N=59,297 |
| --- | --- | --- | --- |
| Age at assessment centre – mean (SD) | 56.7 (8.0) | 57.0 (8.1) | 56.6 (7.9) |
| Highest education attained – % (n) |  |  |  |
| College or university | 39.1 (35,158) | 40.6 (16,697) | 37.9 (18,461) |
| A or AS levels | 13.7 (12,280) | 12.7 (5,216) | 14.5 (7,064) |
| O levels or GCSEs | 26.2 (23,545) | 23.2 (9,553) | 28.7 (13,992) |
| CSEs | 6.5 (5,858) | 6.7 (2,761) | 6.4 (3,097) |
| NVQ | 8.1 (7,249) | 11.2 (4,598) | 5.4 (2,651) |
| Other professional | 6.4 (5,726) | 5.6 (2,283) | 7.1 (3,443) |
| Smoking status – % (n) |  |  |  |
| Never | 53.9 (58,821) | 48.5 (24,282) | 58.5 (34,539) |
| Former | 35.6 (38,885) | 39.4 (19,698) | 32.5 (19,187) |
| Current | 10.5 (11,420) | 12.2 (6,081) | 9.0 (5,339) |
| Alcohol frequency – % (n) |  |  |  |
| Never | 3.1 (3,438) | 1.7 (870) | 4.3 (2,568) |
| Former | 3.5 (3,847) | 3.3 (1,646) | 3.7 (2,201) |
| Special occasions only | 10.6 (11,554) | 6.6 (3,312) | 13.9 (8,242) |
| Once/month to twice/week | 37.6 (41,155) | 35.1 (17,629) | 39.7 (23,526) |
| Thrice/week to daily | 45.2 (49,434) | 53.3 (26,729) | 38.3 (22,705) |
| BMI – mean (SD) | 27.4 (4.8) | 27.8 (4.2) | 27.01 (5.1) |
| Genetic risk score for BMI – mean (SD) | 0.0081 (0.00032) | 0.0096 (0.00054) | 0.0090 (0.00047) |
| WHR – mean (SD) | 0.87 (0.090) | 0.94 (0.065) | 0.82 (0.069) |
| Genetic risk score for WHR – mean (SD) | 0.0082 (0.00038) | 0.0090 (0.00071) | 0.0081 (0.00045) |
| Uses statin drugs – % (n) | 16.4 (17,995) | 21.8 (10,949) | 11.9 (7,046) |

‘Eligible’ defined as having data on genotype and being of white British ethnicity, plus: BMI or WHR, age, sex, genetic PC 1-10, and $\geq 1$ NMR-measured metabolic trait. Genetic risk score values reflect the average per-SNP effect on BMI or WHR.

### MR estimates of the effects of BMI and WHR on metabolic traits measured using NMR

Higher BMI (per SD, or 4.8 kg/m^2^) was estimated to increase concentrations of VLDL particles, but to decrease concentrations of apoB and other lipoprotein types. These estimates were of small magnitude and not greatly altered upon adjustment for WHR, except for VLDL concentration which became negative/inverse (**Figure 1;** full results in **Supp Table 1**). The same pattern was seen for BMI with cholesterol in each particle type, e.g., -0.12 SD (95% CI = -0.15, -0.08) lower LDL cholesterol after adjustment for WHR. In contrast, higher BMI was estimated to increase triglycerides in each lipoprotein type, and these effect estimates were fully attenuated upon adjustment for WHR. Negative effect estimates of BMI with fatty acid ratios and positive effect estimates of BMI with amino acids were each also strongly attenuated with adjustment for WHR (**Figure 2; Supp Table 1**).

**Figure 1.**
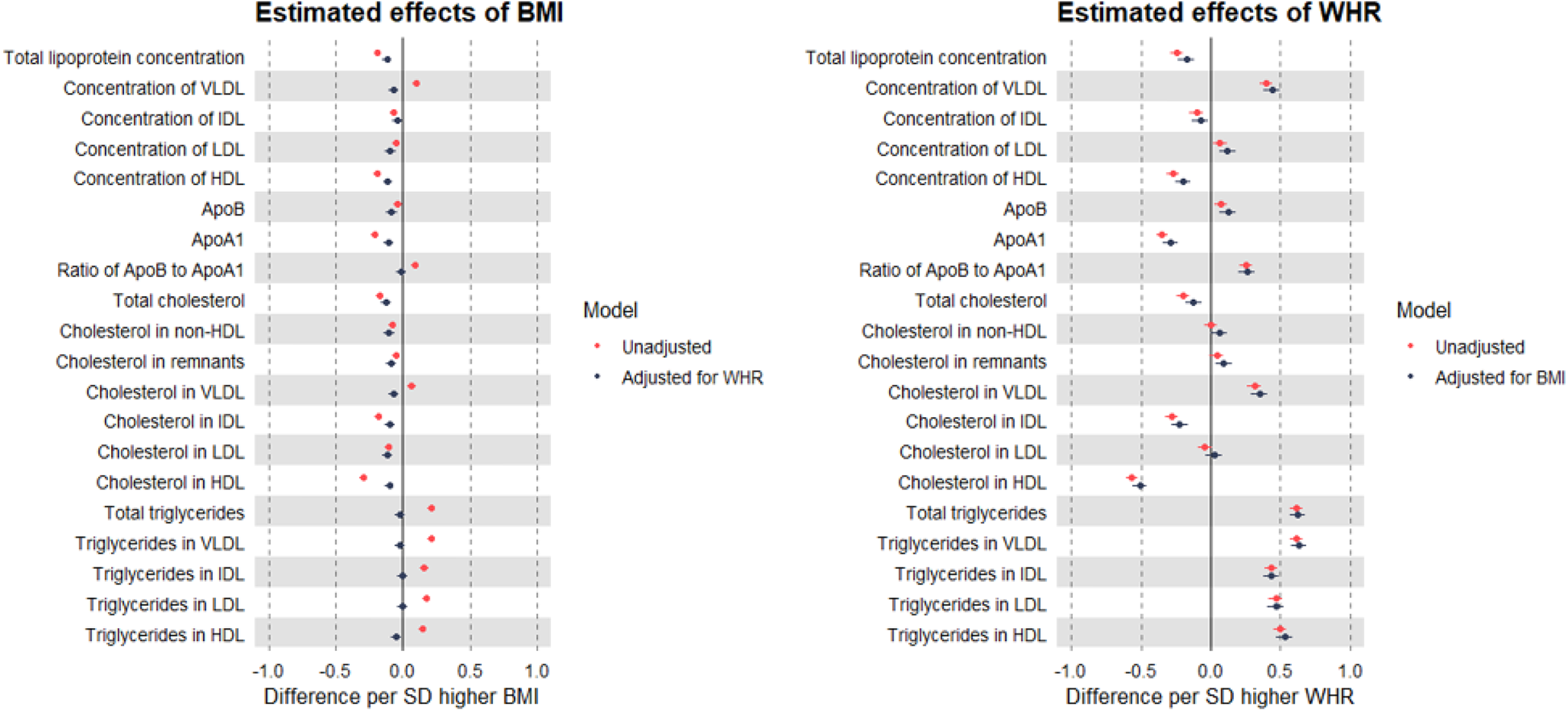
MR estimates of the total (unadjusted) and direct (adjusted) effects of BMI and WHR on lipoproteins, cholesterol, and triglycerides measured using NMR among 109,532 UK Biobank participants Estimates are standardised betas and 95% confidence intervals. ApoB: Apolipoprotein B. ApoA-1: Apolipoprotein A-1. VLDL: Very-low-density lipoprotein. IDL: Intermediate-density lipoprotein. LDL: Low-density lipoprotein. HDL: High-density lipoprotein.

**Figure 2.**
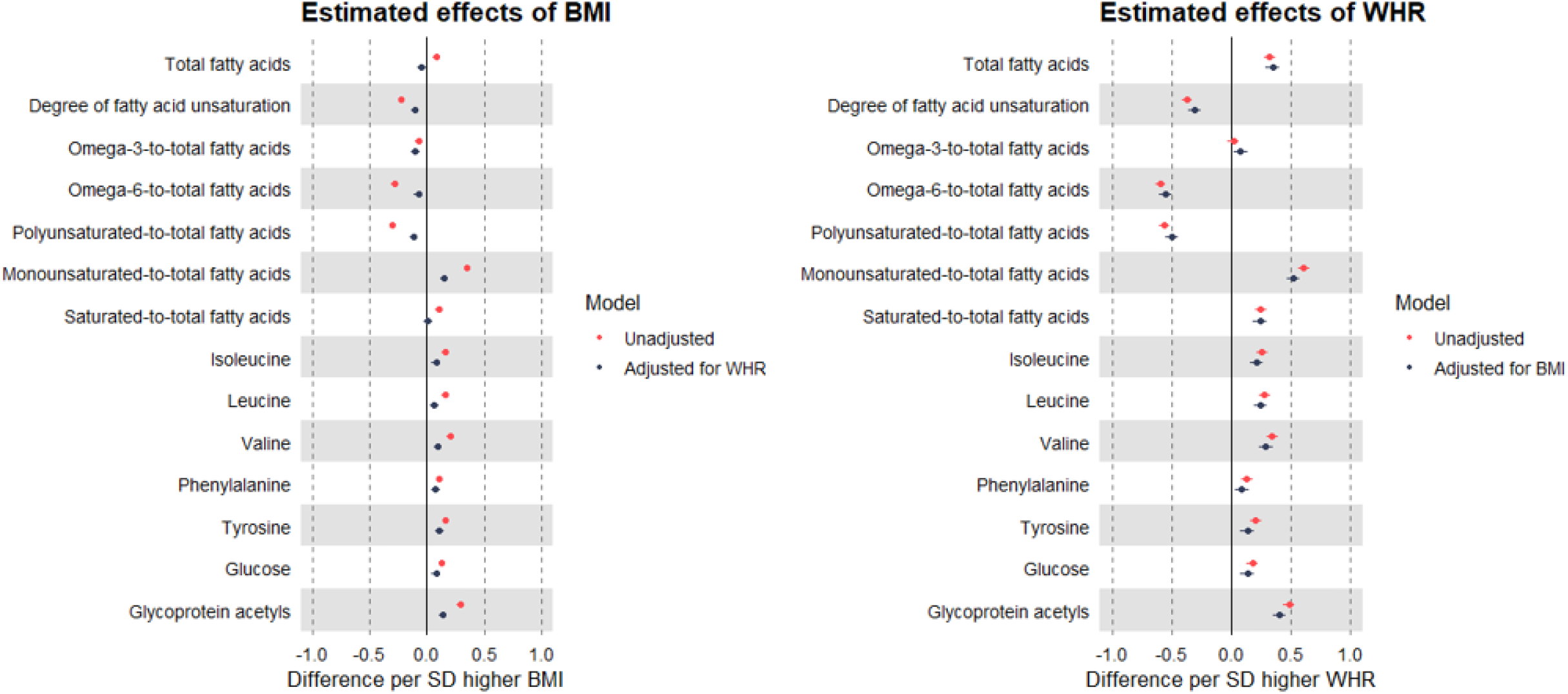
MR estimates of the total (unadjusted) and direct (adjusted) effects of BMI and WHR on selected metabolites measured using NMR among 109,532 UK Biobank participants Estimates are standardised betas and 95% confidence intervals.

Estimated effects of WHR (per SD, or 0.090 ratio-unit) on lipoproteins, lipids, and metabolites were generally of a larger magnitude than seen for BMI, including a positive estimate of effect on apoB, but a small negative-to-null estimate of effect on cholesterol in LDL. Notably, these effect estimates were not altered by adjustment for BMI, with similarly large effects still estimated for triglycerides, e.g., point estimates for VLDL triglycerides were 0.62 and 0.63 SD before and after BMI adjustment, respectively (**Figure 1; Supp Table 1**). This pattern for WHR of larger effect magnitude and lack of attenuation when adjusting for BMI was also seen in relation to numerous metabolites including fatty acid ratios, branched chain amino acids, glucose, and GlycA (**Figure 2**).

The same patterns of effect estimates were seen for metabolic traits including LDL cholesterol based on conventional clinical/biochemistry (non-NMR) measures (**Supp Table 1**).

### Sex-specific MR estimates of the effects of BMI and WHR on metabolic traits measured using NMR

Strong evidence was seen for the interaction of BMI and WHR with sex in relation to most metabolic traits (**Supp Table 2**). Among men, higher BMI (per SD) was estimated to modestly decrease cholesterol in non-HDL particles including LDL, but to increase triglycerides in each lipoprotein type (**Figure 3; Supp Table 3**). Upon adjustment for WHR, effect estimates were moderately attenuated for non-HDL cholesterol, but minimally attenuated for triglycerides. Effect estimates were also negative for BMI with several fatty acid ratios and consistently positive for BMI with amino acids, glucose, and GlycA; none of these estimates attenuated upon adjustment for WHR (**Figure 4; Supp Table 3**). Among women, patterns of estimated effects of BMI on lipids and metabolites were highly consistent with patterns previously seen among sexes combined: higher BMI was estimated to modestly decrease non-HDL particle concentrations and cholesterol, whilst increasing triglycerides. These estimates were largely attenuated with adjustment for WHR, particularly for triglycerides which showed full attenuation (**Figure 3; Supp Table 4**).

**Figure 3.**
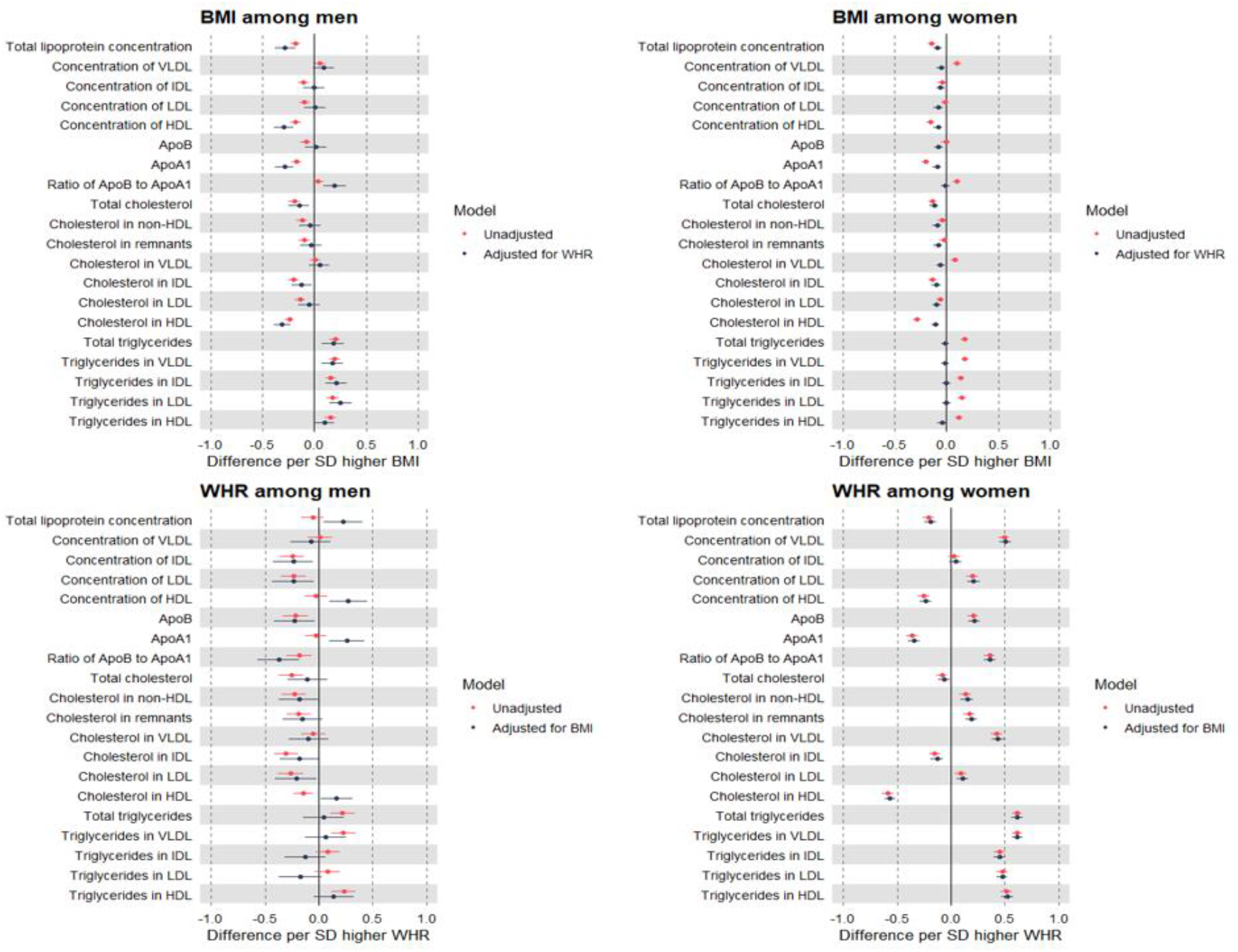
Sex-specific MR estimates of the total (unadjusted) and direct (adjusted) effects of BMI and WHR on lipoproteins, cholesterol, and triglycerides measured using NMR among 109,532 UK Biobank participants Estimates are standardised betas and 95% confidence intervals. ApoB: Apolipoprotein B. ApoA-1: Apolipoprotein A-1. VLDL: Very-low-density lipoprotein. IDL: Intermediate-density lipoprotein. LDL: Low-density lipoprotein. HDL: High-density lipoprotein.

**Figure 4.**
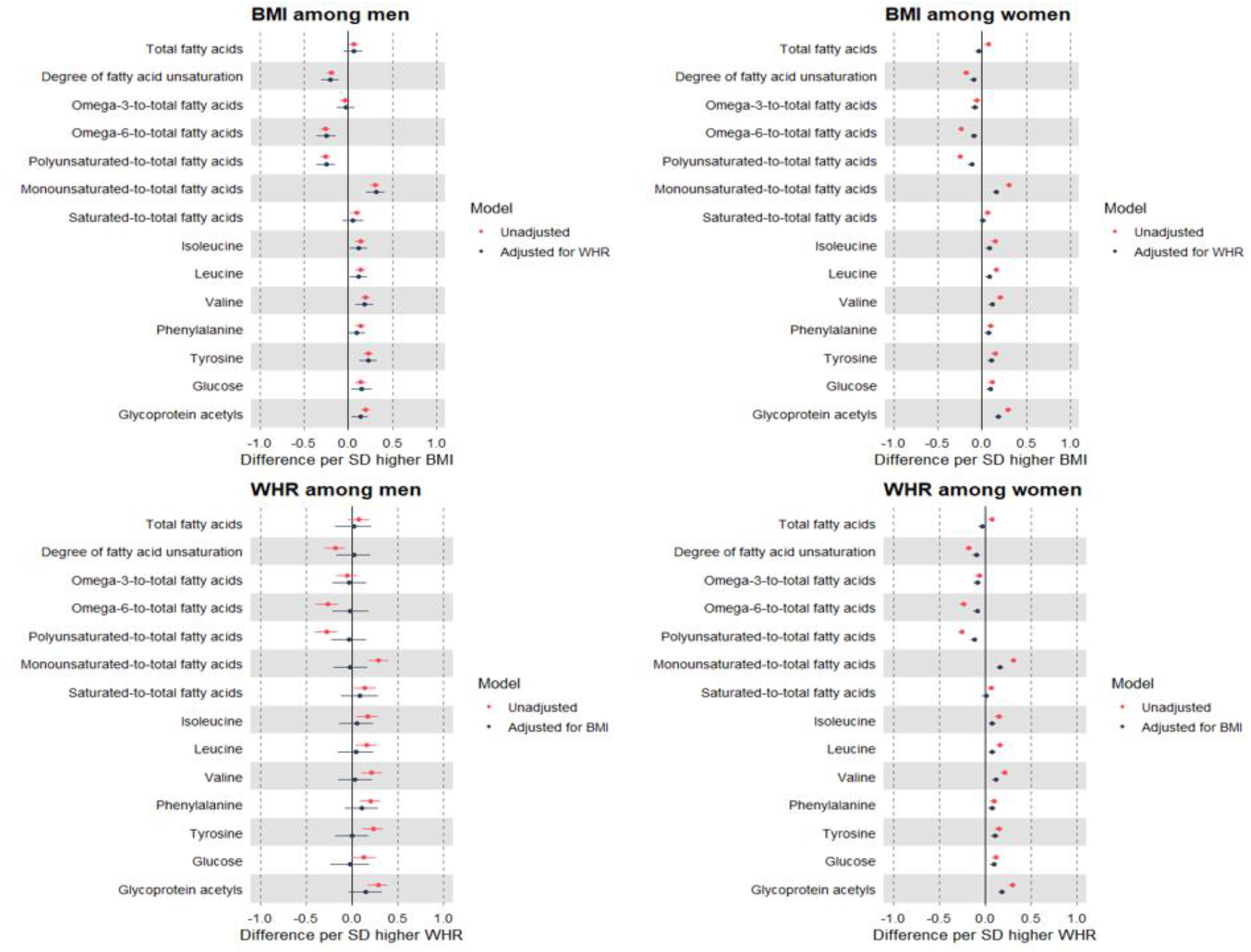
Sex-specific MR estimates of the total (unadjusted) and direct (adjusted) effects of BMI and WHR on selected metabolites measured using NMR among 109,532 UK Biobank participants Estimates are standardised betas and 95% confidence intervals. ApoB: Apolipoprotein B. ApoA1: Apolipoprotein A1. VLDL: Very-low-density lipoprotein. IDL: Intermediate-density lipoprotein. LDL: Low-density lipoprotein. HDL: High-density lipoprotein.

Among men, higher WHR (per SD) was estimated to modestly decrease levels of non-HDL particle concentrations and cholesterol, whilst modestly increasing triglycerides. These effect estimates showed large attenuations upon adjustment for BMI, particularly for non-lipid traits including fatty acids, amino acids, and glucose (**Figures 3-4; Supp Table 3**). Among women, the estimated effects of higher WHR were of a generally larger magnitude, including estimated effects on increasing levels of VLDL concentration, apoB, and triglycerides; none of these effect estimates were attenuated upon adjustment for BMI. Attenuation of WHR effects by adjustment for BMI was more apparent but still modest for metabolites including several fatty acid ratios, branched chain amino acids, and GlycA (**Figure 4; Supp Table 4**).

Sex-specific effect estimates for metabolic traits including LDL cholesterol were again similar when measured using the conventional biochemistry (non-NMR) assay (**Supp Tables 2-4**).

### MR estimates of the effects of BMI and WHR on statin use: sex-combined and sex-specific

Higher BMI (per SD) was estimated to increase the relative odds of using statins by 1.76 (95% CI = 1.68, 1.83) times higher, which increased further when adjusting for WHR (**Supp Table 5**). Higher WHR (per SD) was estimated to increase these odds more strongly, by 2.48 (95% CI = 2.32, 2.66) times higher, which increased further when adjusting for BMI. Effect estimates were also positive within each sex separately, with similar estimates of total and direct effects of BMI among men and women. In contrast, the estimated total and direct effects of WHR on relative odds of using statins were larger among women, e.g., the direct effect estimate was 3.86 (95% CI = 3.74, 3.99) times higher odds versus 2.91 (95% CI = 2.84, 2.99) times higher odds among men.

### MR estimates of the effects of BMI and WHR on metabolic traits measured using NMR, excluding statin users: sex-combined and sex-specific

Evidence was strong for interaction of BMI with statin use in relation to several metabolic traits, including LDL cholesterol (interaction P = 2.6×10^−6^), VLDL lipids, and apoB **(Supp Tables 6)**. Such evidence was weak for HDL cholesterol (interaction P = 0.81). Evidence for interaction was also weak between WHR and statin use for LDL cholesterol (P = 0.36), but stronger for lipids in VLDL, IDL, and HDL. When MR models were repeated among adults who reported not using statins (**Supp Table 7**), higher BMI was still estimated to have a small direct effect on decreasing LDL cholesterol (by -0.07 SD, 95% CI = -0.11, -0.03, adjusted for WHR), whereas higher WHR was still estimated to have larger direct effects on increasing LDL cholesterol (0.14 SD, 95% CI = 0.07, 0.20) and apoB (0.24 SD, 95% CI = 0.18, 0.30).

These statin-exclusion effect estimates differed substantially by sex, with BMI estimated to have a modestly positive direct effect on LDL cholesterol and apoB among men (**Supp Table 8**), yet these were slightly negative among women (**Supp Table 9**). Higher WHR was estimated to substantially decrease LDL cholesterol and apoB among men (**Supp Table 8**), yet substantially increase these trait levels among women (**Supp Table 9**). The large direct effects of WHR on triglyceride levels that were previously estimated also appeared to remain specific to women when excluding statin users.

Sex-combined and sex-specific effect estimates following statin user exclusions were similar for LDL cholesterol based on the conventional clinical/biochemistry assay (**Supp Tables 6-9**).

### MR estimates of the effects of BMI and WHR on metabolic traits measured using NMR, stratified by age group as a proxy for medication use: sex-combined and sex-specific

Evidence was also strong for interaction between adiposity measures and age in relation to most metabolic traits, particularly non-HDL lipids (**Supp Table 10**). Within the youngest age group (38-53 years, where statin use was 5%), higher BMI and WHR were each estimated to modestly increase LDL cholesterol (0.04 SD, 95% CI = -0.01, 0.08 for total effect of BMI and 0.10 SD, 95% CI = 0.02, 0.17 for total effect of WHR). This estimate for BMI fully attenuated, and the estimate for WHR remained unchanged, upon mutual adjustment (**Supp Table 11**). These direct effects on LDL cholesterol were estimated to become more negative for BMI and less positive for WHR in the intermediate age group (54-62 years, statin use 17%), and more so in the oldest age group (63-73 years, statin use 29%) where the direct effects of BMI and WHR on LDL cholesterol had reversed to -0.19 SD (95% CI = -0.27, -0.11) and -0.05 SD (95% CI = -0.16, 0.06; **Figure 5; Supp Tables 11-13**),respectively.

**Figure 5.**
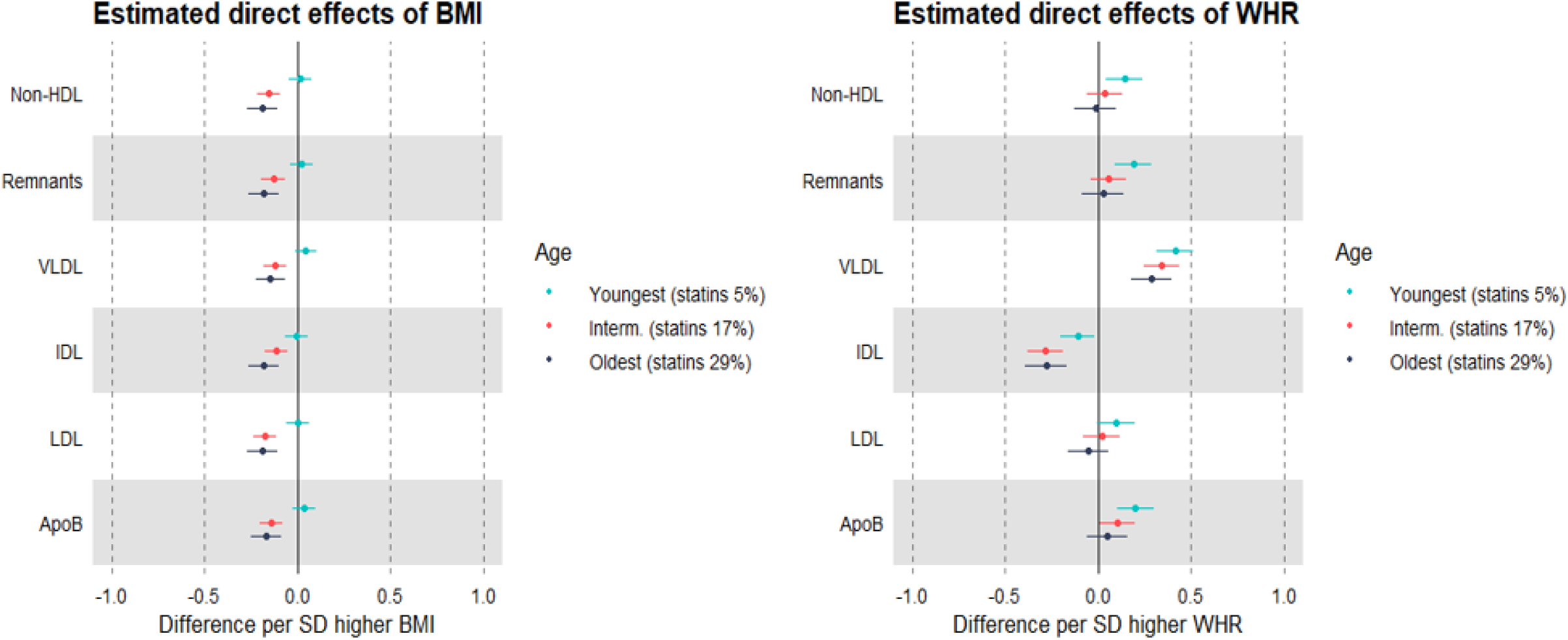
MR estimates of the direct (mutually adjusted) effects of BMI and WHR on non-HDL cholesterol measured using NMR, by age tertile as a proxy for medication use among 109,532 UK Biobank participants Estimates are standardised betas and 95% confidence intervals. HDL: High-density lipoprotein. VLDL: Very-low-density lipoprotein. IDL: Intermediate-density lipoprotein. LDL: Low-density lipoprotein. ApoB: Apolipoprotein B.

Striking differences by sex were also seen in these age stratified estimates of direct effects: among men in the youngest age group (statin use 8%), higher BMI was estimated to directly increase LDL cholesterol, apoB, and other non-HDL traits, e.g. 0.23 SD (95% CI = 0.05, 0.40) for LDL cholesterol (**Figure 6; Supp Tables 14-16**). These effect estimates were directionally reversed among men in the intermediate age group (statin use 23%) and in the oldest age group (statin use 36%), e.g., the LDL cholesterol estimate was -0.24 SD (95% CI = -0.43, -0.05) among the eldest men. In contrast, higher WHR appeared to directly decrease LDL cholesterol in the youngest men, and these estimates appeared null or slightly positive in men of older ages. Among the youngest women, however, the direct effect estimates for BMI were null, and these were negative among women in older age groups where statin use was higher (**Figure 6; Supp Tables 17-19**). Higher WHR was estimated to directly increase LDL cholesterol among the youngest women where statin use was lowest, and these estimates appeared less positive among women in older age groups where statin use was higher (**Supp Tables 14-19**). Estimates for HDL cholesterol, triglycerides, and metabolites appeared to be less affected by age stratifications.

**Figure 6.**
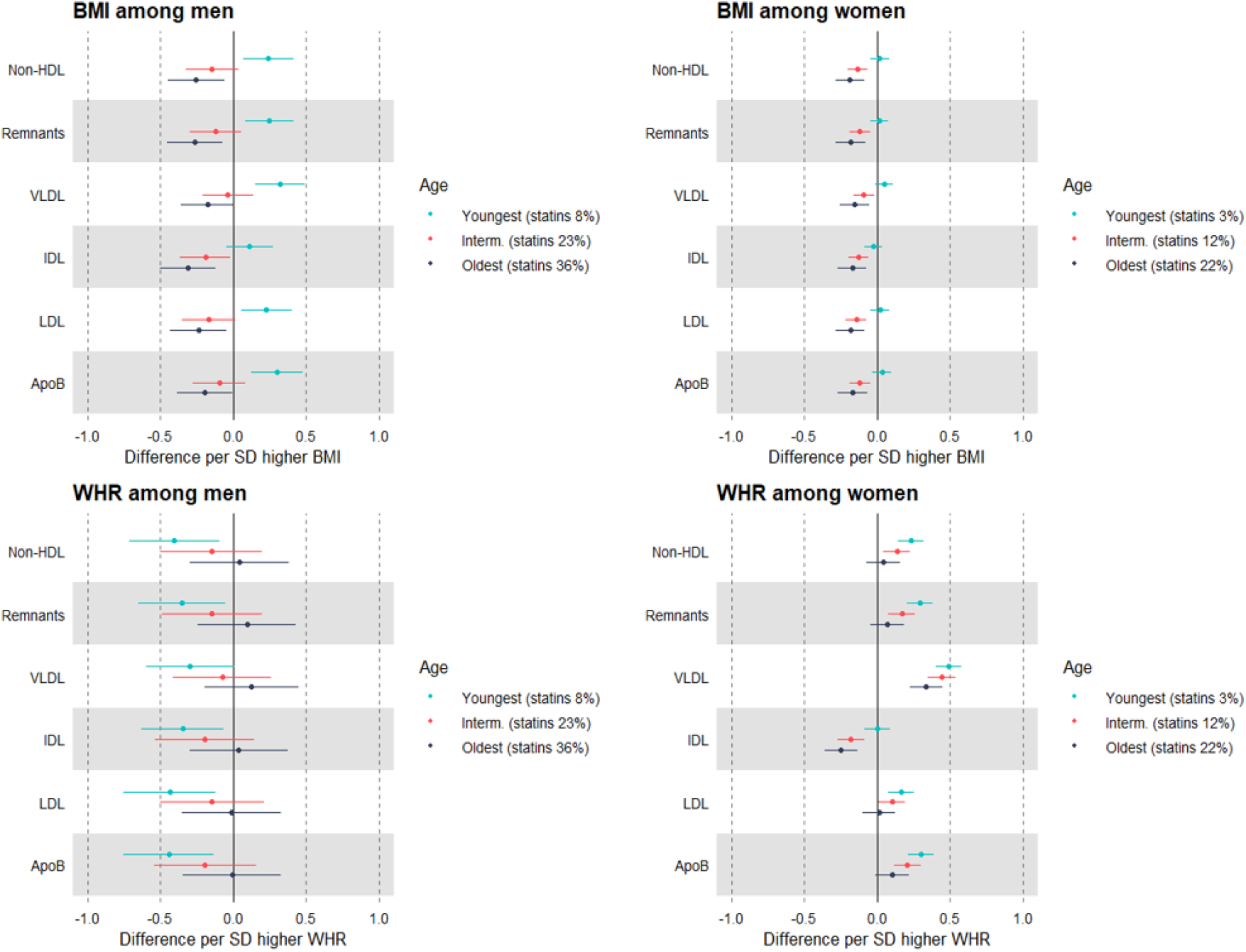
Sex-specific MR estimates of the direct (mutually adjusted) effects of BMI and WHR on non-HDL cholesterol measured using NMR, by age tertile as a proxy for medication use among 109,532 UK Biobank participants Estimates are standardised betas and 95% confidence intervals. HDL: High-density lipoprotein. VLDL: Very-low-density lipoprotein. IDL: Intermediate-density lipoprotein. LDL: Low-density lipoprotein. ApoB: Apolipoprotein B.

Once again, sex-combined and sex-specific effect estimates following age stratifications were similar for LDL cholesterol based on conventional biochemistry measures (**Supp Tables 10-19**).

### Comparison of effect estimates using conventional observational models

Higher BMI (per SD) was estimated to increase levels of numerous lipid traits, most notably higher concentrations of, and lipids in, VLDL particles and their subclasses, and higher triglycerides, LDL cholesterol, and apoB (**Supp Figures 1-2; Supp Table 20**). These estimates showed modest attenuation upon adjustment for WHR, with attenuations appearing greatest for triglycerides. Effect estimates for WHR were of a similar pattern but often larger magnitude, including more positive effect estimates with triglycerides which showed modest attenuation when adjusting for BMI. Evidence was strong for interaction of BMI and WHR with sex in relation to most metabolic traits (**Supp Table 21**). Among men, effect estimates indicated that higher BMI increased VLDL lipids and triglycerides, but modestly lowered LDL cholesterol which remained apparent when adjusting for WHR (−0.05 SD, 95% CI = -0.06, -0.04; **Supp Figures 3-4; Supp Table 22**). Higher WHR was estimated to slightly increase LDL cholesterol (0.03 SD, 95% CI = 0.01, 0.04 adjusted for BMI). Mutually adjusted estimates among men tended to be larger for BMI than for WHR, particularly for fatty acid ratios and amino acids. Among women, BMI and WHR were each estimated to increase LDL cholesterol and apoB, but with magnitudes tending to be higher for WHR, particularly in relation to triglycerides, fatty acid ratios, and GlycA, which were robust to adjustment for BMI (**Supp Figures 3-4; Supp Table 23**).

Consistent with MR results, higher BMI (per SD) was estimated to increase the relative odds of using statins by 1.61 (95% CI = 1.60, 1.63) times higher, which attenuated to 1.40 (95% CI = 1.39, 1.42) times higher when adjusting for WHR (**Supp Table 24**). These estimates were higher for WHR, particularly among women. Evidence was strong for interaction of BMI and WHR with statin use in relation to most metabolic traits including LDL cholesterol (interaction P < 6.7×10^−20^; **Supp Table 25**). When excluding statin users, BMI and WHR were each estimated to raise LDL cholesterol (mutually adjusted estimates for BMI and WHR were 0.04 SD (95% CI = 0.03, 0.05) and 0.12 SD (95% CI = 0.11, 0.13), respectively), and similarly for apoB (**Supp Table 26**). These were more pronounced among women than men, particularly for WHR (**Supp Tables 27-28**). Evidence was also strong for interaction of BMI and WHR with age in relation to most metabolic traits, and more so for

LDL than for HDL traits (**Supp Table 30**). Within the youngest age group (statin use 5%), higher BMI and WHR were each estimated to increase LDL cholesterol before mutual adjustment (0.08 SD, 95% CI = 0.07, 0.09 for BMI and 0.16 SD, 95% CI = 0.15, 0.18 for WHR) and both estimates remained positive after mutual adjustment, with a larger estimate for WHR at 0.14 SD (95% CI = 0.13, 0.16; **Supp Figure 5; Supp Table 31**). These estimates for LDL cholesterol became negative with BMI and less positive with WHR at intermediate ages (statin use 17%), and more so in the oldest age group (statin use 29%; **Supp Figure 5; Supp Tables 32-33**). These were more pronounced among women, particularly for WHR (**Supp Figure 6; Supp Tables 34-39**). These patterns were similar for other LDL-related traits including apoB, whilst estimates for HDL cholesterol and metabolites were less affected by statin user exclusions and age stratifications.

The same pattern of results was seen for biochemistry-derived (non-NMR) metabolic traits including LDL cholesterol (**Supp Tables 20-39**).

## Discussion

In this study, we aimed to better compare the direct/isolated effects of total and abdominal adiposity on markers of systemic metabolism. We used new high-throughput metabolic data on ∼110,000 adults from the UK Biobank, representing at least a five-fold increase in sample size over previous studies (19, 21, 30, 46), and we applied both MR and conventional observational approaches to enable more robust causal inference. Our results suggest that abdominal adiposity has a dominant role in driving the metabolic harms of excess adiposity – by raising harmful lipoprotein, lipid, and metabolite levels – particularly among women. Such direct impact of abdominal adiposity may be underestimated by conventional observational approaches. Our findings also suggest that apparent effects of adiposity on lowering LDL cholesterol are explained by an effect of adiposity on statin use.

We estimated causal effects using one-sample MR in individual-level data which uses GRSs as instruments and should be less prone to confounding and reverse causation bias than conventional multivariable regression models. MR evidence was strongest for an adverse total (unadjusted) effect of WHR on most metabolic traits, also seen with conventional models, in contrast to more comparable estimates seen between BMI and WHR in previous studies with much smaller sample sizes (19, 21, 46). Current patterns of attenuation upon mutual adjustment for adiposity measures also differed greatly by method. MR estimates supported almost complete attenuation of the total effects of BMI when adjusting for WHR, most notably for lipoprotein triglycerides. These attenuations of the total effects of BMI when adjusting for WHR were only modest based on conventional models.

In some cases, the direct effects of BMI (on inclusion of WHR in multivariable MR) appeared to be slightly beneficial for non-HDL lipids, suggesting a relatively harmless role of peripheral adiposity. This supports earlier genetic findings of greater insulin sensitivity from higher adiposity stored in body limb and gluteofemoral compartments (47, 48), and findings on ‘favourable adiposity’ alleles which raise body fat percentage whilst improving metabolic risk factor profiles (47, 49, 50).

These support the theory of impaired expandability of peripheral adipose tissue which results in lipid ‘spill over’ into visceral and ectopic compartments and ultimately metabolic dysfunction (51-53); this process may be proxied by direct effects of higher BMI (total adiposity) which do not operate through WHR (abdominal adiposity). Likewise, whilst conventional observational estimates suggested that WHR only partially alters lipids and glycemic and inflammatory metabolites, MR estimates supported larger total effects of WHR and further suggested that these effects are largely independent of BMI, i.e., there was no attenuation in multivariable MR models. Together, this suggests that conventional observational methods may underestimate the direct metabolic impact of abdominal adiposity. This was especially apparent for triglycerides which appeared exclusively tied to WHR based on MR.

Our results also suggest that men and women do not experience the metabolic harms of adiposity equally. Among men, MR estimates suggested that BMI has a dominant role in most metabolic traits, with minimal attenuation of BMI effects upon adjustment for WHR (including those effects on triglycerides), yet nearly full attenuation of WHR effects upon adjustment for BMI. Among women, however, MR estimates suggested a dominant role of WHR in these effects with virtually no attenuation upon adjustment for BMI, yet nearly full attenuation of BMI effects upon adjustment for WHR. This dominant pattern for WHR with lipids and metabolites closely resembled the pattern seen for WHR among sexes combined. Men are recognised to typically store more fat abdominally, potentially as a result of a lower capacity for peripheral adipose tissue to expand (52). WHR is expected to proxy impaired adipose expandability among both sexes, but higher WHR may better mark the ‘extremeness’ of impaired expandability among women since higher WHR would only be expected when peripheral compartments are greatly overwhelmed. The apparent exclusivity of these direct effects of WHR to women is unexpected, however. This could be a reflection of stronger genetic instruments among women (particularly for WHR) rather than genuine biological differences, or sex-specific selection biases, e.g., higher BMI is known to reduce the likelihood of participating in population-based studies and genetic estimates from UK Biobank suggest that this likelihood is lower for women (54). Representative samples would be valuable for replicating these findings.

Total and abdominal adiposity are expected to be co-dependent, such that one influences the other. It is thus important not to dismiss total effects of BMI in either sex since attenuation here would likely reflect mediation rather than confounding. BMI has proven useful as a non-invasive and cost-effective indirect measure of total adiposity which correlates highly (r > 0.8) with total fat mass as well as trunk fat mass measured using DXA scans, and which generates similar estimates of effect (as DXA measures) on markers of systemic metabolism in childhood, young adulthood, and middle adulthood (21, 30, 55). Such ‘BMI effects’ are expected to be underpinned by a higher volume of adiposity stored abdominally, viscerally, and ectopically.

LDL cholesterol is an established cause of CHD (13), but whether adiposity raises LDL cholesterol has been surprisingly unclear. Four previous MR studies based mostly on indirectly measured LDL cholesterol found either slightly positive, slightly inverse, or null effects of BMI (6, 23, 56, 57), yet a moderately positive effect of WHR-adjusted-for-BMI in the one study examining this (6). In contrast, two other MR studies based on LDL cholesterol measured directly by NMR spectroscopy supported a positive effect of BMI (19, 46) (WHR was not examined). Our current MR estimates suggested a small inverse effect of BMI on LDL cholesterol among men and women, yet a small positive effect of WHR among women only. These inconsistencies may be partly attributable to statin use, which was common in our sample (22% of men and 12% of women). We also found MR evidence that BMI and WHR each directly raise the relative odds of using statins. Since statins lower LDL cholesterol (13), this could explain apparent ‘protective effects’ of adiposity on LDL cholesterol.

Indeed, we found strong evidence that the effects of BMI and WHR on numerous lipids and metabolites differed by statin use, more so for LDL and VLDL than for HDL cholesterol, as expected. When repeating analyses with statin users excluded, MR estimates of BMI (total and direct) appeared to remain negative/inverse and WHR appeared to be positive. This analysis could be prone to collider bias, however, since statin use could be a mediator between adiposity and metabolites (or a common effect of adiposity and metabolites), and stratifying on colliders can induce non-causal associations via mediator-outcome confounding or selection-induced correlations (58). Age, in contrast, would not be a collider since it is not influenced by adiposity, and age may proxy for broad exposure to statins (and other medications) since their use increases markedly with age. We found strong evidence of interaction by age to support this, and, when stratifying analyses of adiposity and metabolic traits by age tertile, we found that apparent effects of higher adiposity on lowering LDL cholesterol and other non-HDL traits were specific to older ages where statin use was most common (29% in the oldest group). BMI and WHR each raised LDL cholesterol and other non-HDL traits including apoB at the youngest ages when statin use was rare (5% in the youngest group), with direct effects appearing specific to WHR. These findings, together with those of adiposity influencing statin use, strongly suggest that apparent effects of adiposity on lowering LDL cholesterol are explained by medication use. They also highlight the challenge of isolating the effects of exposures which influence the use of common medications, even with MR. This could help explain inconsistencies in results for adiposity and conventionally-measured LDL cholesterol across previous MR studies, which typically included adult samples and did not interrogate the role of statins or other medications (6, 23, 56, 57). Stratifying by age in such instances may be one simple approach to reveal and overcome these distortions.

### Study limitations

We used WHR as an indirect measure of abdominal adiposity because this is feasibly measured at large scale and has an available set of strong genetic instruments, but this is less precise than DXA or more advanced body scanning devices. WHR is likely preferable to WC as an indicator of abdominal fatness since WHR shares a weaker phenotypic correlation with BMI than does WC (25). Another notable limitation of this study is the unrepresentative nature of UK Biobank (response rate ∼5% (33). The distribution of BMI in UK Biobank closely matches its distribution in the more representative Health Survey for England, however, and the majority of effect estimates for common lifestyle risk factors with CVD mortality do appear consistent in terms of direction and magnitude (59). The impact of selection bias may be greatest for socioeconomic and behavioural risk factors which are prone to high measurement error and instability over time (60); these should be less problematic for adiposity exposures. In practice, robust conclusions and recommendations do not come from a single cohort study but rather are assembled via consistency of results from across multiple cohorts and, importantly, from across multiple study designs which carry distinct sources of bias (61). We encourage such triangulation of evidence using observational and MR estimates (62), as done previously for BMI and metabolites (19) and for statins and metabolites (31), which can in turn be compared with randomised controlled trial estimates. It would be valuable to re-estimate the effects investigated here in other settings, in diverse ancestries, and with other approaches. It would also be valuable when data exist at scale to examine more precise adiposity measures in relation to metabolomic and proteomic traits to better characterise disease susceptibility.

## Conclusions

The results of this large MR study using new high-throughput metabolic data from UK Biobank suggest that abdominal adiposity has a dominant role in driving the metabolic harms of excess adiposity – by raising harmful lipoprotein, lipid, and metabolite levels – particularly among women. Such direct impact of abdominal adiposity may be underestimated by conventional observational methods. These findings also suggest that apparent effects of adiposity on lowering LDL cholesterol are explained by an effect of adiposity on statin use.

## Data Availability

Individual-level data are available via application to UK Biobank (managed access).

## Acknowledgements

The authors are grateful to the UK Biobank participants and study team for access to data to undertake this study (Project #30418).

## Data availability

Individual-level data are available via application to UK Biobank (managed access).

## Author contributions

JAB planned the study, conducted analyses, and wrote the first draft. HJ, AC and PW contributed to the generation and curation of the NMR metabolic data. All others critically reviewed the intellectual content of manuscript drafts and with JAB approved the final version for submission.

