## Supplementary Figures for "Dominant role of abdominal adiposity in circulating lipoprotein, lipid, and metabolite levels in UK Biobank: Mendelian randomization study"

**Supplementary Figure 1** Conventional observational estimates of the total (unadjusted) and direct (adjusted) effects of BMI and WHR on lipoproteins, cholesterol, and triglycerides measured using NMR among 89,540 UK Biobank participants

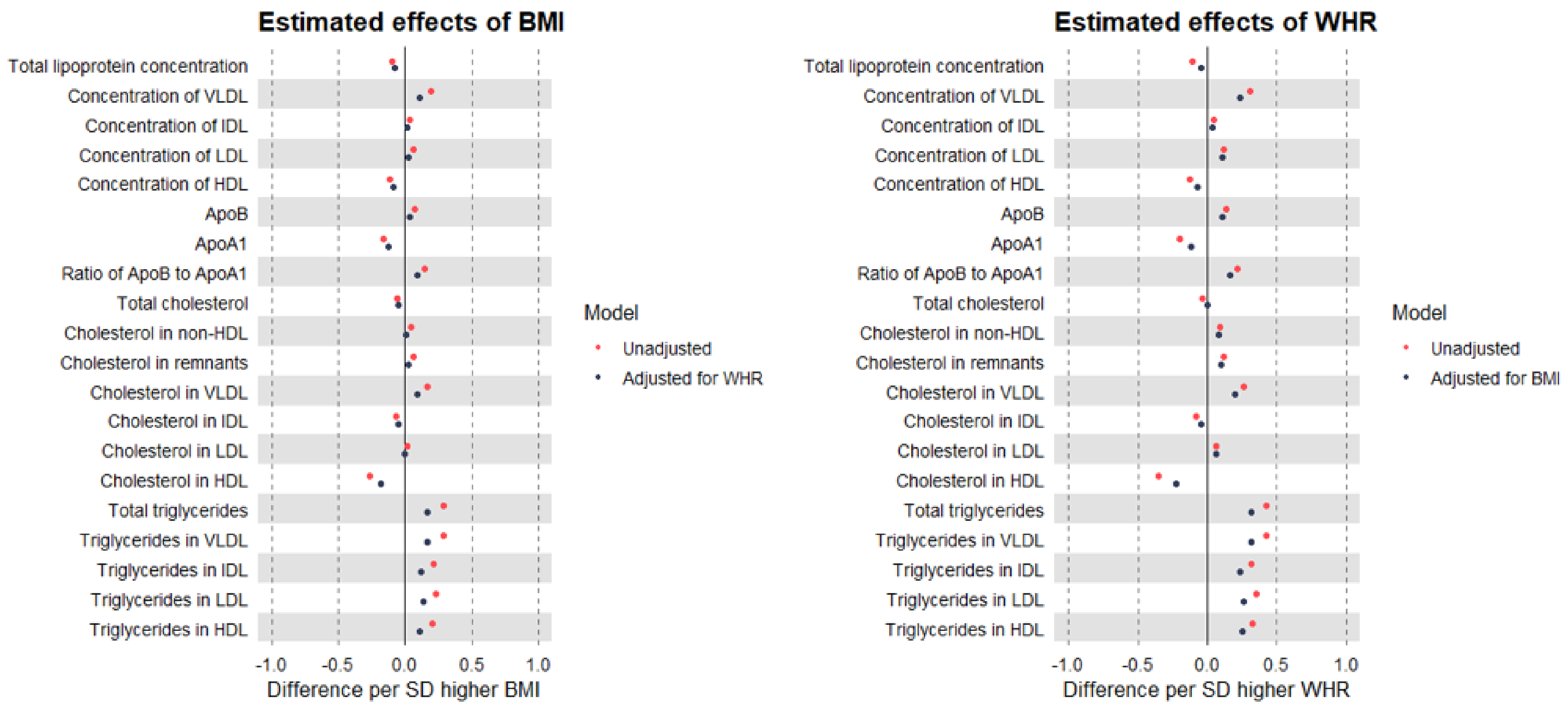


Estimates are standardised betas and 95% confidence intervals (not visible due to small standard errors). ApoB: Apolipoprotein B. ApoA-1: Apolipoprotein A-1. VLDL: Very-low-density lipoprotein. IDL: Intermediate-density lipoprotein. LDL: Low-density lipoprotein. HDL: High-density lipoprotein.

**Supplementary Figure 2** Conventional observational estimates of the total (unadjusted) and direct (adjusted) effects of BMI and WHR on selected metabolites measured using NMR among 89,540 UK Biobank participants

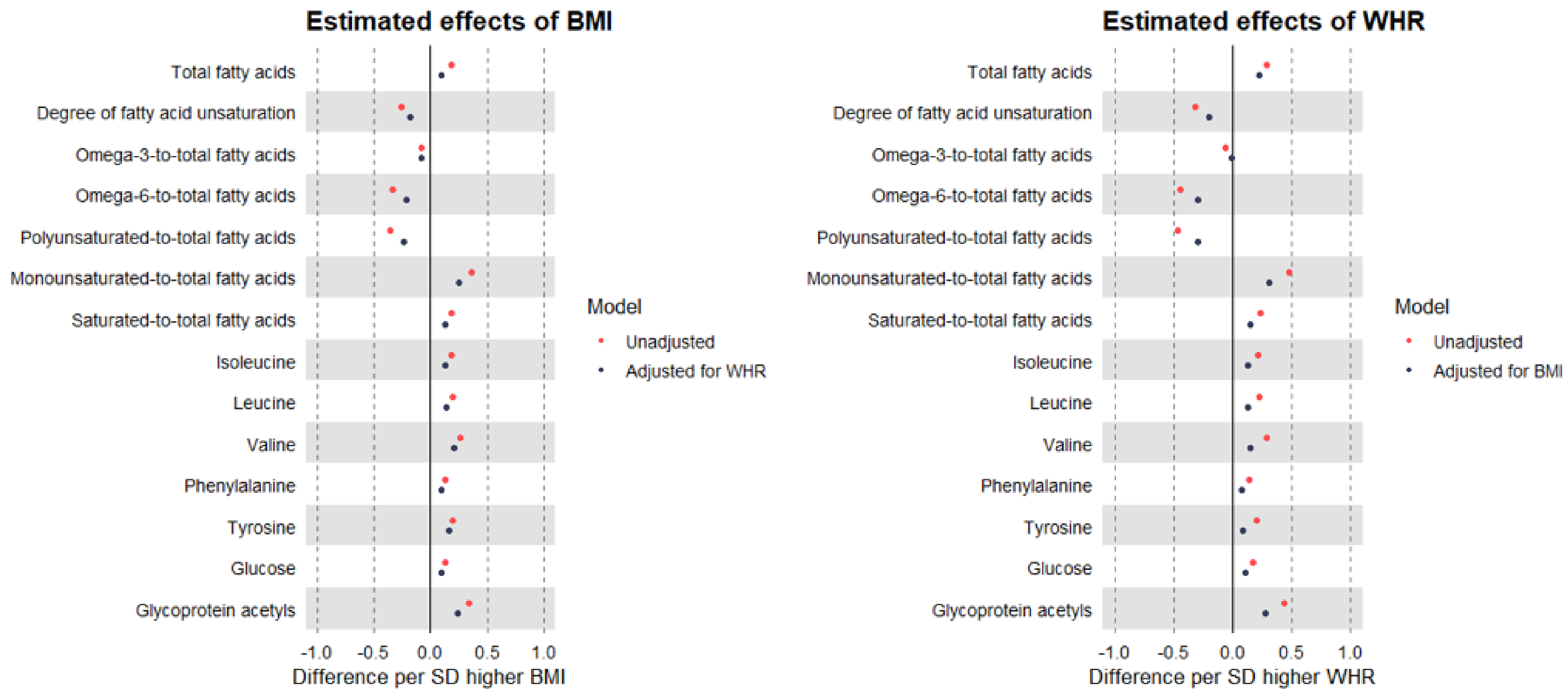


Estimates are standardised betas and 95% confidence intervals (not visible due to small standard errors).

**Supplementary Figure 3** Sex-specific conventional observational estimates of the total (unadjusted) and direct (adjusted) effects of BMI and WHR on lipoproteins, cholesterol, and triglycerides measured using NMR among 89,540 UK Biobank participants

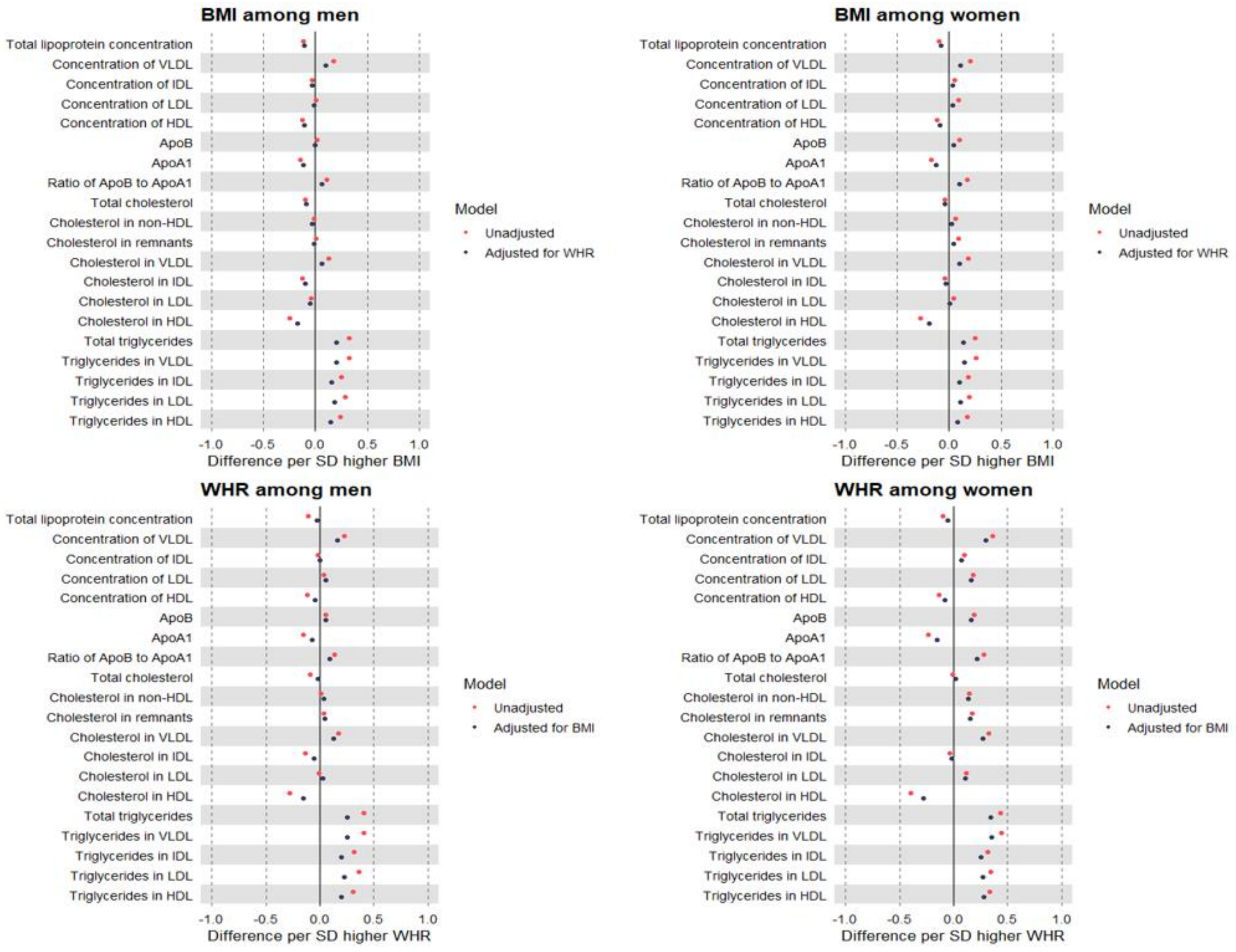


Estimates are standardised betas and 95% confidence intervals (not visible due to small standard errors). ApoB: Apolipoprotein B. ApoA-1: Apolipoprotein A-1. VLDL: Very-low-density lipoprotein. IDL: Intermediate-density lipoprotein. LDL: Low-density lipoprotein. HDL: High-density lipoprotein.

**Supplementary Figure 4** Sex-specific conventional observational estimates of the total (unadjusted) and direct (adjusted) effects of BMI and WHR on selected metabolites measured using NMR, among 89,540 UK Biobank participants

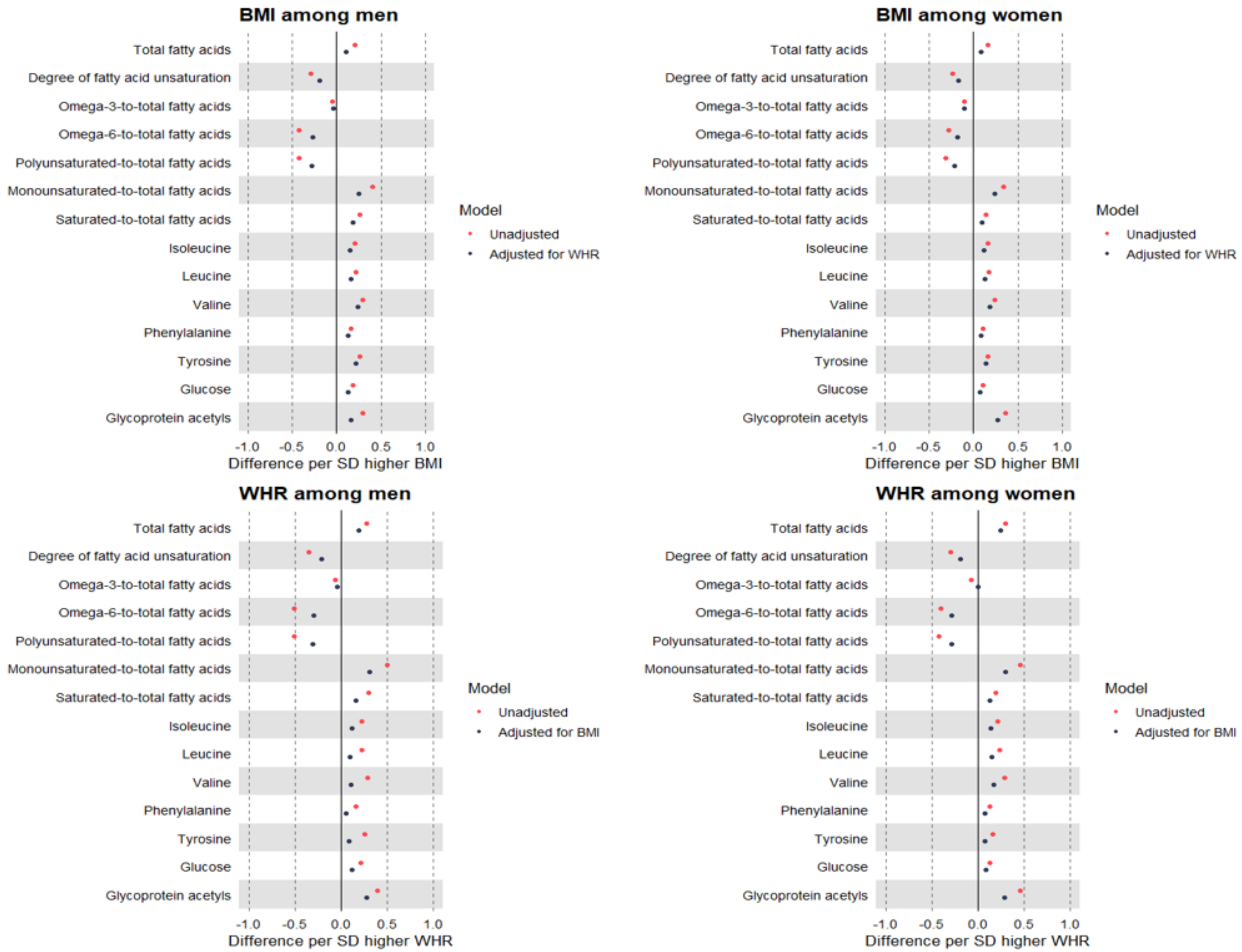


Estimates are standardised betas and 95% confidence intervals (not visible due to small standard errors). ApoB: Apolipoprotein B. ApoA1: Apolipoprotein A1. VLDL: Very-low-density lipoprotein. IDL: Intermediate-density lipoprotein. LDL: Low-density lipoprotein. HDL: High-density lipoprotein.

**Supplementary Figure 5** Conventional observational estimates of the direct (mutually adjusted) effects of BMI and WHR on non-HDL cholesterol measured using NMR, by age tertile as a proxy for medication use among 89,540 UK Biobank participants

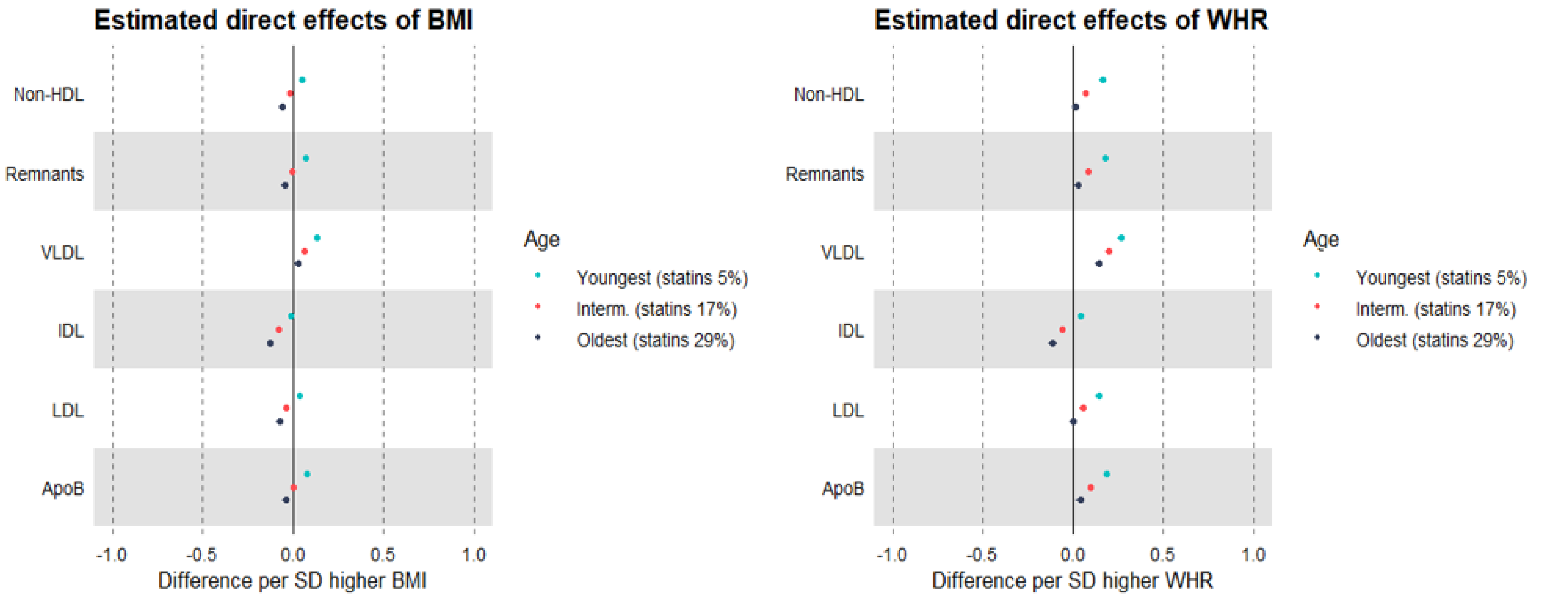


Estimates are standardised betas and 95% confidence intervals (not visible due to small standard errors). HDL: High-density lipoprotein. VLDL: Very-low-density lipoprotein. IDL: Intermediate-density lipoprotein. LDL: Low-density lipoprotein. ApoB: Apolipoprotein B.

**Supplementary Figure 6** Sex-specific conventional observational estimates of the direct (mutually adjusted) effects of BMI and WHR on non-HDL cholesterol measured using NMR, by age tertile as a proxy for medication use among 89,540 UK Biobank participants

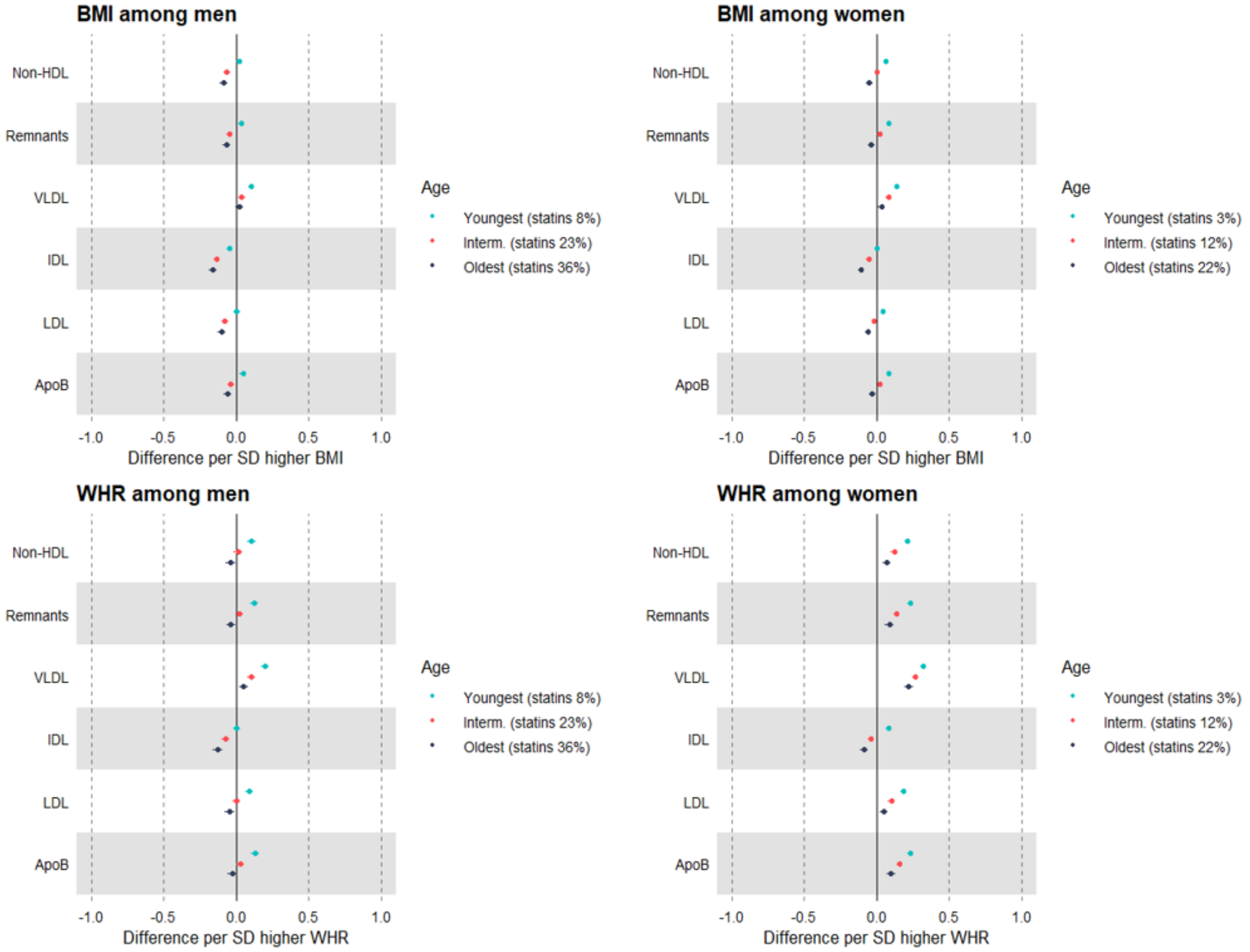


Estimates are standardised betas and 95% confidence intervals (not visible due to small standard errors). HDL: High-density lipoprotein. VLDL: Very-low-density lipoprotein. IDL: Intermediate-density lipoprotein. LDL: Low-density lipoprotein. ApoB: Apolipoprotein B.
